# Mortality prediction by Carotid Intimal Medial Thickness with a novel Life Table Statistical Model

**DOI:** 10.1101/2025.02.03.25321583

**Authors:** Christopher M Rembold

## Abstract

Carotid Intimal Medial Thickness (CIMT) is an ultrasound estimate of preclinical atherosclerosis. The Charlottesville CIMT trial entered 1470 subjects from 1995 to 2011 and assessed total mortality in 2024 with a mean 15-year follow-up. In those subjects who reported no smoking or coronary disease, Cox statistics revealed that the presence of plaque and 4^th^ quartile common, bulb and internal CIMT measurements predicted total mortality (compared to the 1^st^ quartile). Age-normalized CIMT quartiles were less predictive. The age of subjects in the 4^th^ quartile was 15 years higher than the 1^st^ quartile. To correct for different ages, a novel life table model was created to analyze clinical data from a cohort who entered a trial at varying ages. This life table model allowed inclusion of subjects lost to follow-up. With this life table model, age-normalized 4^th^ and 3^rd^ quartile common, bulb and internal CIMT predicted total mortality. These data show the utility of measuring CIMT to predict mortality and the benefit of the life table model. The output statistic of the life table model, Age Difference, is an easily understandable and relevant measure of effect size.

## INTRODUCTION

In people without known atherosclerotic disease (primary prevention), it is complicated to decide who would benefit from treating dyslipidemia to prevent coronary artery disease (CAD), myocardial infarction (MI) and other atherosclerotic diseases. The most common clinical approach is to treat based on low density lipoprotein (LDL) levels (1). Unfortunately, there are other predictors of atherosclerotic outcomes such as high density lipoprotein (HDL) levels, lipoprotein little a (Lp_a_), family history, smoking, systolic blood pressure (SBP), diabetes, exposure to small airborne particles (PM2.5) and a sedentary lifestyle (1). There are people with a high LDL and a negative family history that live a long life without atherosclerotic complications. There are also people with a low LDL who have a MI at a young age.

One approach to deciding who to treat is computed tomography (CT) coronary calcium scoring which has been shown to predict atherosclerotic complications quite well in older people (2). Calcium scoring has little operator variability. Unfortunately, in the MESA trial a woman has to be 66 and a man has to be 52 for each to have a 50% chance of a positive calcium score; so younger people have less predictive accuracy (2).

Carotid Intimal Medial Thickness (CIMT) is a high-resolution ultrasound of the carotid arteries performed to predict atherosclerosis burden and therefore risk (3). The carotids are close enough to the skin for more accurate ultrasound measurement than coronary arteries. Both thickness and plaque can be measured. Unfortunately, CIMT measurements have issues with operator variability: proper imaging requires more operator time and therefore cost. Also, some labs look only at common CIMT while others (like ours) look at common, bulb and internal CIMT. In a meta-analysis, common CIMT was only modestly predictive of atherosclerotic outcomes (4).

In 1995, the University of Virginia started a clinical CIMT program. We did a retrospective analysis in 2004 (the published Charlottesville CIMT trial (5)) and found that CIMT predicted major cardiac outcomes (MI, stroke (CVA), transient ischemic attack (TIA) or revascularization). The odds ratio for being in the 4^th^ (top) quartile of age-normalized carotid bulb CIMT was 5.8 more that the 1^st^ (lowest) quartile and the p for quartile trend was 0.007. Internal age-normalized CIMT was also predictive, the p for quartile trend was 0.03. The common carotid CIMT was not significantly predictive.

In 2024, total mortality was evaluated in the Charlottesville CIMT trial. Interestingly, traditional Cox proportional hazard statistics did not predict outcomes well for such a prolonged trial with subjects entering at varying ages, so I created a new life table model. This model and its benefits will be discussed in detail.

## METHODS

### CLINICAL TRIAL

All CIMT studies performed by the cardiovascular division at the University of Virginia between 11/29/1995 and 1/11/2011 were included in this analysis – see reference (5) for details including the imaging protocol. The program was begun on 11/29/1995 and had two intermediate analyses, one in 2004 (the published trial (5)) and the other in 2011 (similar results yet not accepted for publication despite multiple attempts). A quality assurance database has been in place. Vital status (dead vs. alive vs. lost to follow-up) was ascertained in 2023 via medical record search by the author from January to March 2024. This quality assurance database was then converted into a research database in April 2024. The protocol was approved by the University of Virginia Human Investigation Committee (#10915).

### NOVEL LIFE TABLE MODEL

Data was arranged in a spreadsheet with a row for each subject: column A was status at end of the trial (0=alive, 1=dead), column B was age at start of study, column C was age at end of study if alive or the age of death, column D was years of data collected, column E was missing years (0 if alive at study closure), and columns F and beyond were 0 or 1 for various sorting groups.

Data was imported into Matlab (Natick, MA, USA). Data were first tested with a Cox proportional hazard model that did not include those subjects lost to follow-up (6, 7). This was done in Matlab with [b,logl,H,stats] = coxphfit(group (column F+), data duration(0,column D), ‘Censoring’, datcensoring’ (column A)).

The novel models were based on a life table with a row for each subject and a column for each age. Fig. 1 shows the data from the first 17 nonsmokers in the clinical database. Subjects 1-15 were patients who did not die during the duration of the trial while subjects 16 and 17 died during the trial.

**Figure 1.**
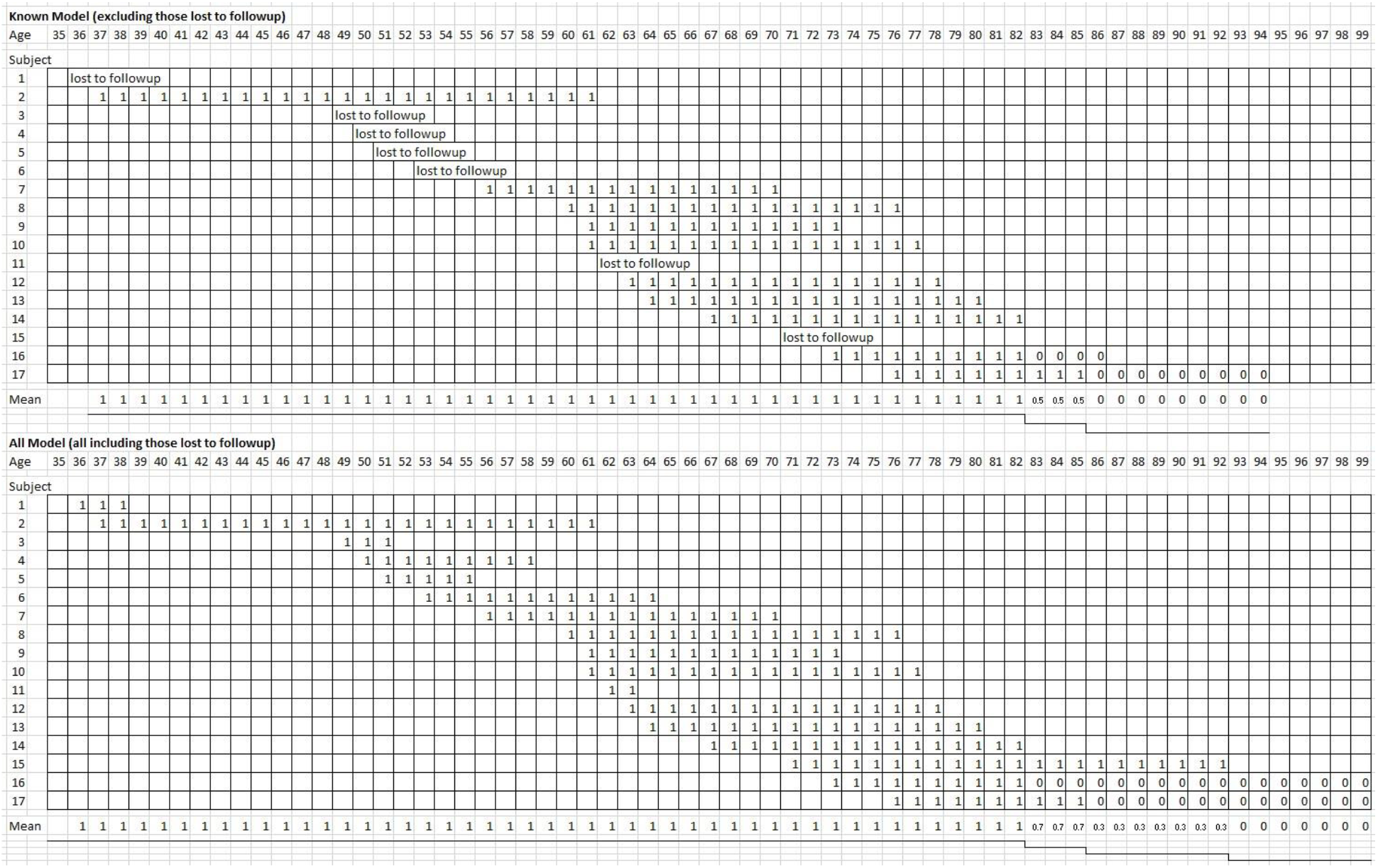
Example of the two life table models with the first 17 no smoking subjects. ONEs are entered for the age when subjects are known to be alive and ZEROs for ages when subjects are dead.

The life table model called KNOWN was conservative like the Cox model because those subjects lost to follow-up were not included (Fig. 1, top). ONES were entered for every age when a subject was known to be alive for the entire trial duration. If a subject died during the trial, ONES were entered for the ages when the subject was alive and ZEROS were entered from the age of death until the trial ended. Subjects 1, 3, 4, 5, 6, 11 and 15 were lost to follow-up and are not included. Subject 16 and 17 died at 83 and 86 and the ONES prior were replaced by ZEROS until the end of the trial. The deaths at age 83 and 86 cause the mean to drop to 50% then 0. The end of data occurred in 2024 when subject 17 had been dead for 9 years and the data were collected.

The life table model called ALL also included those subjects lost to follow-up (Fig. 1, bottom: see the results and discussion section for rationale). ONES for every age when a subject was known to be alive for at least 2 years. Once a subject was lost to follow-up or the trial ended, the ONES were no longer included. If a subject died during the trial, ONES were entered for the ages when the subject was alive and ZEROS were entered from the age of death until age 105. Subject 1 and 3 were lost to follow-up and are only included for short time. Subjects 16 and 17 died at 83 and 86 and the ONES prior were replaced by ZEROS causing the mean to drop to 67% then 33% at their death ages (see line at bottom). The end of data for subject 15 who was lost to follow-up at age 92 caused the mean to drop to 0% until age 105.

Data was analyzed for the KNOWN and ALL life table models as the Age Difference (AD) between groups. In columns F and beyond, subjects were divided into two groups based on various subject characteristic, e.g. smoking. The mean life table data for mortality for the two groups was plotted (Fig. 2). As is clearly visible, the curves appeared to be approximately sigmoid and did not fit a typical proportional hazard plot, so hazards were not calculated. The mean life table data was divided into 20 quantiles (see horizontal lines in Fig. 2 separating the mortality into 20 groups: the first (top) was from 1.0 to 0.95 shown in light blue, and the eleventh from 0.5 to 0.45 shown in grey). Statistics were then performed only those quantiles in which each group had at least a minimum number of subjects (this prevented rare events from dominating statistics - for this analysis the minimum n was 8). The age difference (AD) between the two groups was calculated from the area enclosed within each quantile that had enough n. In Fig. 2, 11 quantiles (colored horizontal stripes) fit this criterion. Then mean AD (mAD), standard error AD (seAD = standard deviation/square root of n of quantiles), Z for AD (zAD = mAD/seAD), and p value for 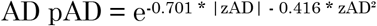 (value limited between 1 and 0.000001) were then calculated. Matlab code is included in the appendix.

**Figure 2.**
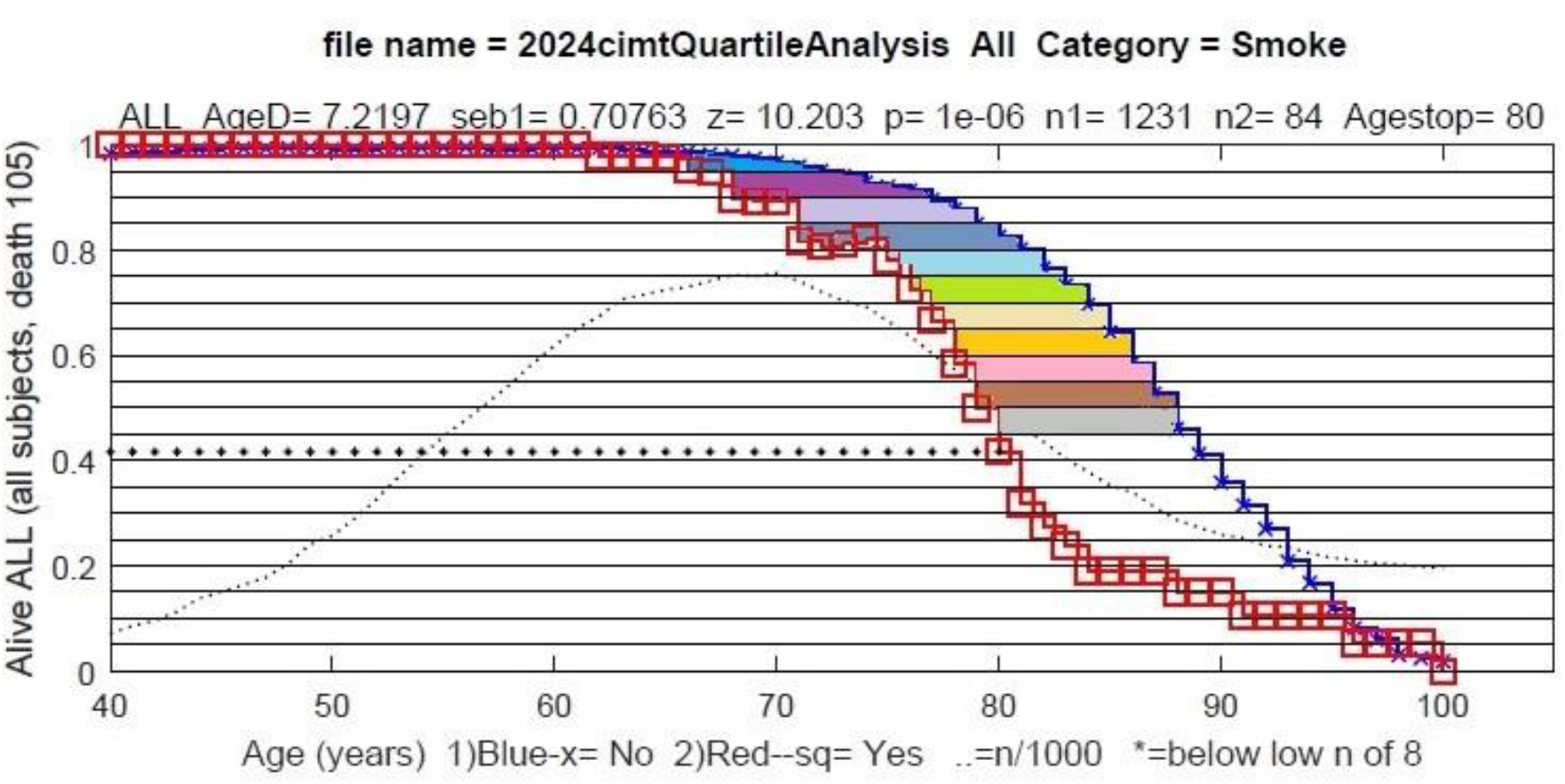
The method to calculate Age Difference. The mean number alive from the whole CIMT cohort separated by those not smoking (blue x) and those smoking (red □) groups. Horizontal lines were drawn at 0.05 intervals creating 20 quantiles (horizontal stripes). The area between the red and blue lines represents the age difference (AD) for each quantile (area was calculated vertically given that the life table data did not always decrease as subjects came and went in the life table - see red line in dark blue quantile at ages 70-75). The top 11 quantiles were colored in as they fit the criterion that there were at least a certain number of subjects in each of the groups (in this case beyond age 80 there were less than 8 subjects in the red group – the line of asterisks show where data were analyzed). The mean AD (in this case 7.2 years) for each of these top 11 quantiles were calculated and statistics performed (shown just above the graph). The dotted line is the total number of subjects/1000 at each age

## RESULTS

There were 1470 subjects studied: 671 were men and 799 women. There were 194 subjects (13%) who were known to have died, 663 alive (45%) at the analysis date (2023), 613 lost to follow-up (42%). The mean follow-up was 15 years (range 0-27) and the average number of missing years of data was 5.7. For those whose status was known at the end of the study, the mean follow-up was 21 years. At the date of the CIMT measurement, the mean age was 56 (range 16-85), 2% reported known CAD, 2% reported a prior CVA or TIA, 4% reported diabetes, 41% reported hypertension, 99% reported dyslipidemia, 7% were smokers, and 31% reported hormone use. The average height was 67 inches (range 56-78), weight 170 pounds (range 70-372), BMI 26.3 (range 13-49), Systolic BP 136 (range 90-204), and Diastolic BP 83 (range 30-120). The median common IMT was 0.83 mm (range 0.4-2.5), bulb IMT 1.2 mm (range 0.45-4.7), internal IMT 0.80 (range 0.3-4.7), and mean Flow Mediate Dilation 16.4% (range -100 to 120).

For the whole cohort, standard Cox statistics revealed that age, male sex, reported CAD, hypertension, smoking and prior stroke or transient ischemic attack (CVA/TIA) predicted mortality (Table 1). Since screening for preclinical atherosclerosis would typically be not performed in those who smoked or had known CAD, data from these two groups (9% of total subjects) were excluded from further analysis. In the no smoking/no CAD cohort, Cox statistics revealed similar results: age, male sex, hypertension and prior CVA/TIA predicted mortality.

**Table 1:**
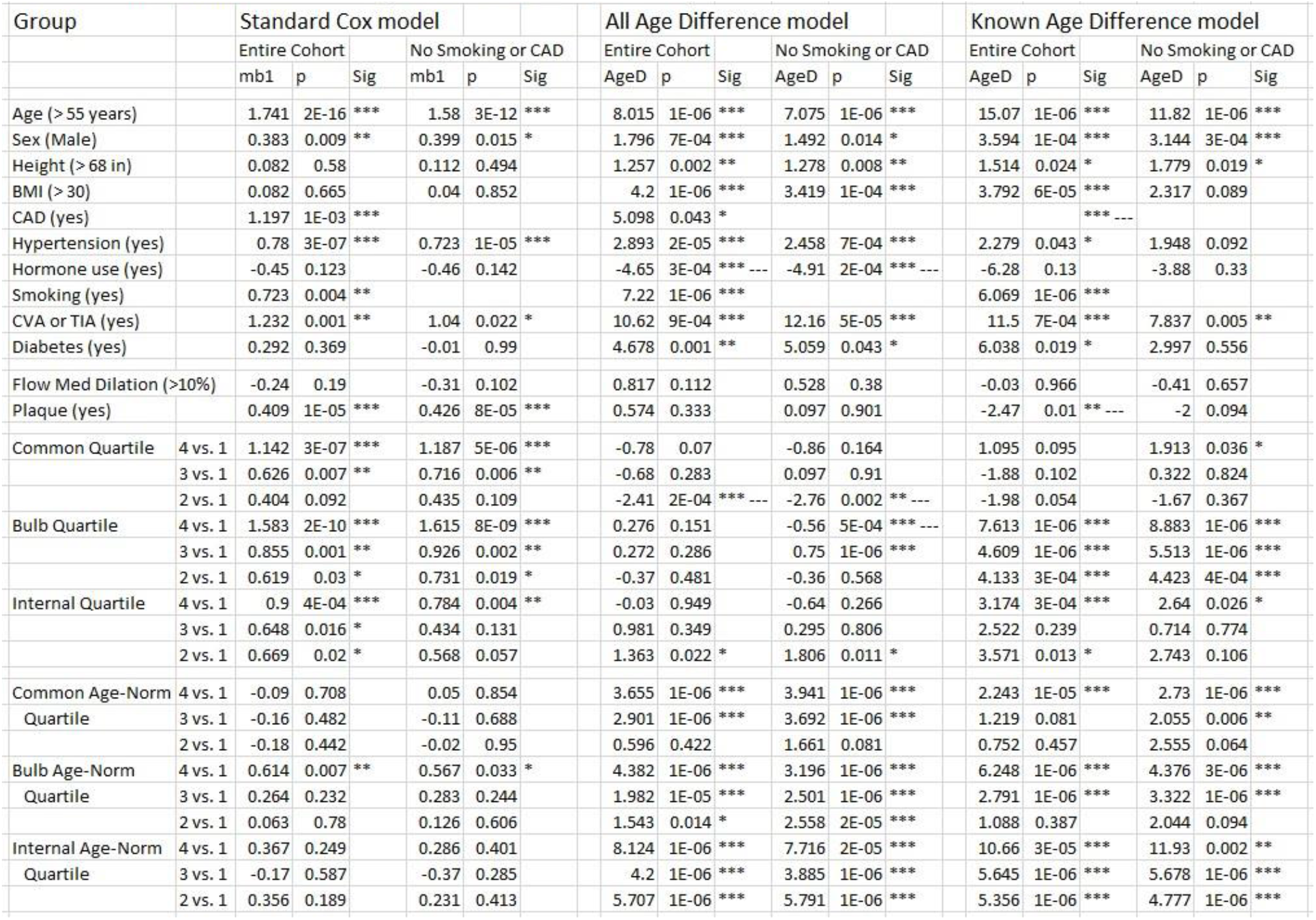
Life Table data for the entire cohort and those who did not smoke or have CAD with the three models. mb1 is the mean hazard ratio for the Cox model, AgeD (AD) is the effect size in years for the life table models. p is the p value, Sig refers to significance (*** is <0.001, ** is < 0.01, * is <0.05 and --- indicates a lower risk).

In the no smoking/no CAD cohort, Cox statistics showed the presence of plaque predicted mortality (Table 1). Flow mediated dilation did not predict mortality. Common (4^th^ and 3^rd^), bulb (4^th^, 3^rd^ and 2^nd^) and internal (4^th^) CIMT measurements predicted total mortality (when compared to the 1^st^ quartile).

In our prior paper with a shorter follow-up duration, we created age-normalized CIMT quartiles that predicted outcomes (5). With Cox statistics, only the bulb (4^th^) age-normalized CIMT predicted total mortality (when compared to the 1^st^ quartile). For the Cox analysis of measured common CIMT, the mean age was 63 in the 4^th^ quartile and the mean age was 48 in the 1^st^ quartile. This age difference and the lack of mortality prediction by age-normalized CIMT quartiles suggested that analyzing a cohort with widely varying ages with Cox statistics (based on time in the study) may not appropriate, perhaps a life table method would be better. So, I created two novel life table models to analyze clinical data from a cohort with a wide age range and a relatively low outcome probability.

The life table models are shown for the first 17 subjects in Fig. 1 (described more fully in methods). Data are placed in two different life tables: 1) The life table model termed “Known” was more conservative and did not include those subjects lost to follow-up (similar to the cox model): 1) for those alive to the end of the trial, a row with ONES (1) were entered for the ages known to be alive and 2) for those who died, ONES would be entered for the ages known to be alive and ZEROES (0) would be entered for the ages known to be dead with a maximal age of the end of the trial (Fig. 1, top). 2) The life table model termed “ALL” was a more inclusive and included all subjects including those lost to follow-up: 1) for those who did not die during the trial, a row with ONES (1) were entered for the ages known to be alive and 2) for those who died, ONES would be entered for the ages known to be alive and ZEROES (0) would be entered for the ages known to be dead with a maximal age of 105 (i.e. beyond the end of the trial, Fig. 1, bottom). The columns were then averaged to create a mean life table for each group with each model. Statistics on the Age Difference (AD) between the two groups was calculated as in the methods (see Fig. 2). AD is an easily understood measure of effect size.

In the no smoking, no CAD group, the Known model (no subjects lost to follow similar to the Cox model) revealed that age, male sex and prior CVA/TIA predicted mortality. Common (4^th^), bulb (4^th^, 3^rd^ and 2^nd^) and internal (4^th^) CIMT measurements predicted total mortality (when compared to the 1^st^ quartile). Age-normalized common (4^th^ and 3^rd^), bulb (4^th^ and 3^rd^) and internal (4^th^, 3^rd^ and 2^nd^) CIMT predicted total mortality. The Known model was similar to the Cox model for CIMT measurements and predicted mortality better than the Cox model for age normalized CIMT quartiles.

Subjects lost to follow-up are usually not included in proportional hazard models because they are more likely to have had an outcome that is not measured (7). Some authors (8) suggest that such subjects can be sometimes be included. I think that the life table model proposed is such a model because the vital status *at each age* entered is known for all of the subjects regardless of follow-up status. Data ends in the life table for both those lost to follow-up and those alive at the end of the trial. Following is the analysis for the All life table model which included subjects lost to follow up.

In the no smoking, no CAD group, the All model (all subjects) revealed that age, male sex, BMI, hypertension, prior CVA/TIA and diabetes predicted mortality (hormone use negatively predicted mortality). Plaque did not predict total mortality. Bulb (4^th^) negatively and (3^rd^) positively predicted total mortality. Age-normalized common (4^th^ and 3^rd^), bulb (4^th^, 3^rd^, and 2^nd^) and internal (4^th^, 3^rd^ and 2^nd^) CIMT predicted total mortality (when compared to the 1^st^ quartile). The All model predicted mortality better than the Cox model for age normalized quartiles.

These results are better shown in the output of the Matlab program (Fig. 3). The left panel compares the 4^th^ vs 1^st^ quartile for measured common CIMT. Cox statistics (top panel) reveal significantly higher hazards for the 4^th^ quartile (red), the All model (center panel) shows no significant difference and the Known model showed a significant AD of 1.9 years. This is likely caused by a higher mean age of 63 in the 4^th^ quartile compared to the age of 48 in the 1^st^ quartile for measured common CIMT. The right panel of Fig. 3 compares the 4^th^ vs 1^st^ quartile for age-normalized common CIMT. The mean age was 57 in the 4^th^ quartile compared to the age of 55 in the 1^st^ quartile for age-normalized common CIMT. Cox statistics (top panel) reveal no significant difference while both the All and Known life table models showed significantly higher hazards for the 4^th^ quartile (red) with AD values of 3.9 and 2.7 years.

**Figure 3.**
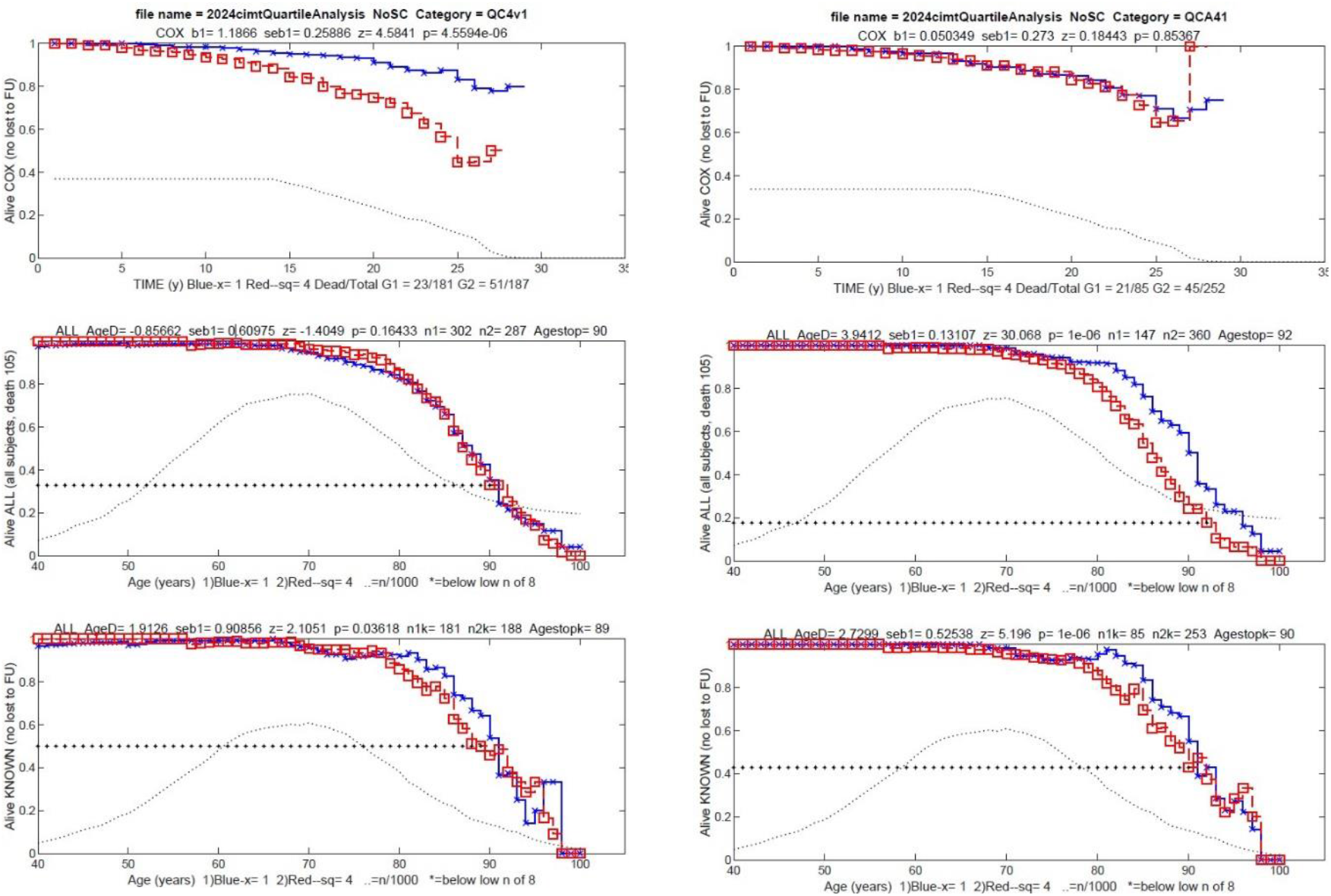
Left: the analysis of measured Common CIMT 4^th^ (red) vs 1^st^ (blue) quartiles. Right: the analysis of Age-Normalized Common CIMT 4^th^ (red) vs 1^st^ (blue) quartiles. The top panels are standard Cox statistics. The center panels are the life table ALL model and the lower panels are the life table KNOWN model. The dotted line in all panels shows the number of subjects/1000 at each age. The asterisks show the lower limit for data with an adequate number of subjects for statistical testing.

Results by age were analyzed by splitting the no smoking, no CAD group cohort by median age at CIMT measurement into two groups: younger ≤ 55 and older ≥ 56 years old (Table 2). In the Cox model, hypertension was predictive at both ages, BMI at lower ages, and male sex, prior CVA/TIA and plaque at older ages. Bulb 3^rd^ quartile CIMT measurements predicted mortality at younger age and bulb and internal 4^th^ quartile at older age. Age normalized CIMT quartiles only predicted mortality in the bulb 4^th^ quartile at older age.

**Table 2:**
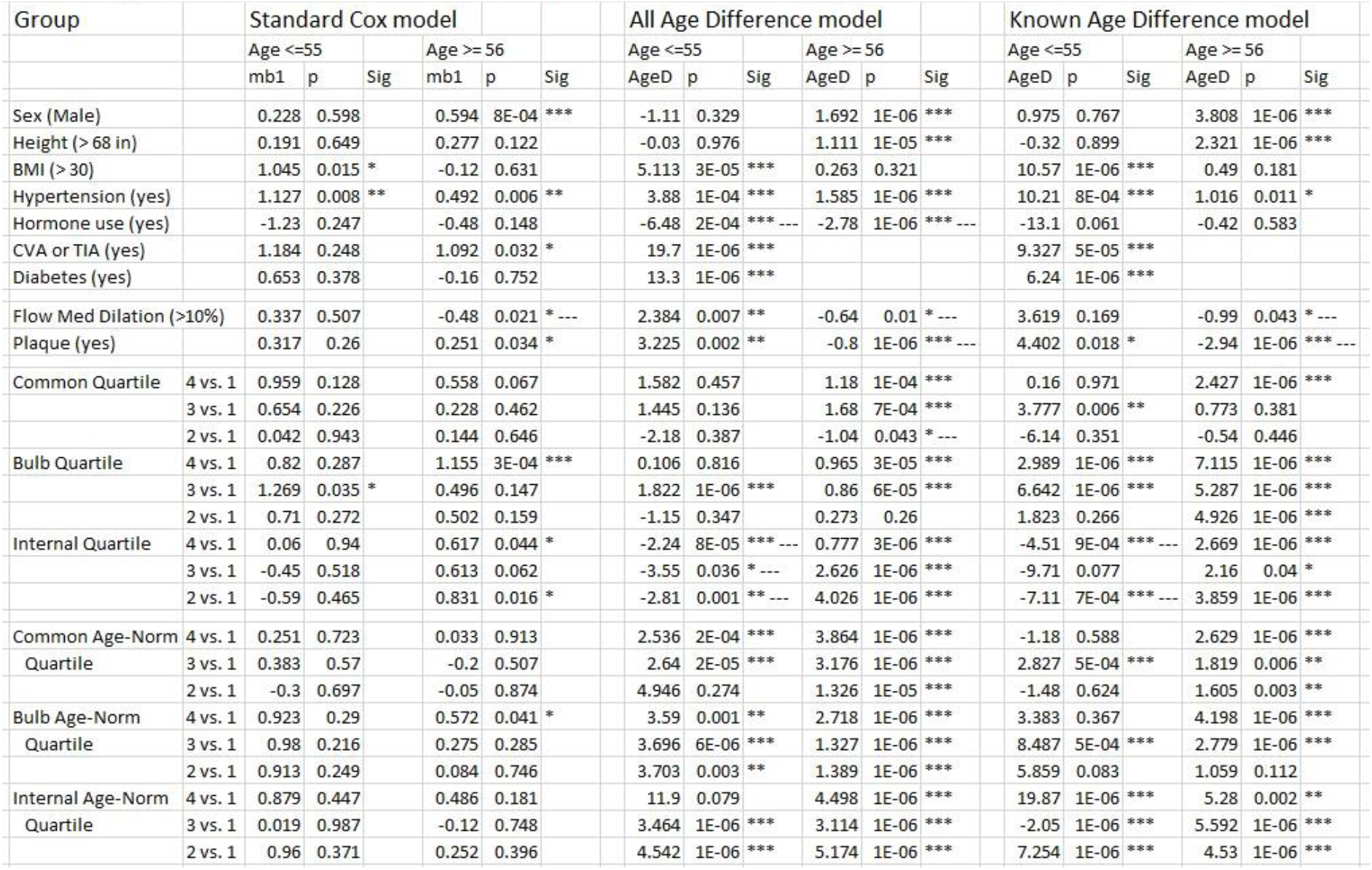
Data for subjects sorted by age younger ≤ 55 and older ≥ 56 at CIMT measurement (all in the no smoking or no CAD cohort. mb1 is the mean hazard ratio for the Cox model, AgeD (AD) is the effect size in years of the life table models. p is the p value, Sig refers to significance (*** is <0.001, ** is < 0.01, * is <0.05 and -- - indicates a lower risk).

In the All model, hypertension was predictive at both ages, BMI, prior CVA/TIA, diabetes, plaque and Flow mediated dilation at lower ages, and male sex at older ages. Hormone use at both ages negatively predicted mortality. Age normalized common, bulb and internal CIMT quartiles mortality at both younger (4^th^ and 3^rd^ quartiles) and older age (4^th^, 3^rd^ and 2^nd^ quartiles). CIMT measurements were less predictive. The Known model had similar results.

## DISCUSSION

### UTILITY OF CIMT TO PREDICT TOTAL MORTALITY

This study shows the benefit of ultrasound screening for carotid atherosclerosis. Magnussen et al (9) report from an analysis of 1.5m people a population attributable risk of 21% for total mortality given 5 modifiable atherosclerotic risk factors of SBP, BMI, non HDL cholesterol (a proxy for LDL cholesterol), smoking and diabetes. With a smaller cohort, this study found that imaged atherosclerosis assessed as age normalized quartiles predicted total mortality. Total mortality is less biased than softer endpoints of MI or cardiovascular death. These data suggest a benefit of atherosclerosis screening with CIMT, especially for younger people who have yet to calcify their coronary arteries (women under age 66 and men under age 52 (2)). This study showed benefit of age normalized CIMT quartiles in younger and older subjects (≤ 55 vs. ≥ 56 years).

Interestingly, when corrected for age, CIMT measurements were less predictive and the presence of plaque was not predictive for total mortality. This shows the importance of considering age (age normalized CIMT quartiles) when predicting outcomes of a chronic condition like atherosclerosis.

Age-normalized thickness was predictive of mortality in the Common, Bulb and Internal Carotids (Tables 1 and 2), both at younger and older ages. This differs from our prior analysis (5), in which age-normalized thickness in the Bulb was the most predictive of MI, stroke and revascularization.

CIMT imaging has some disadvantages (3). There is operator variability. It is typically more expensive than CT coronary calcium scoring. CIMT imaging has the disadvantage of looking at the carotids and not the coronaries. Transthoracic ultrasound has yet to have the imaging capacity to image coronaries.

The clinical trial has some limitations. Entry was based on clinicians ordering the CIMT study, so the population could be biased. The trial was retrospective. Lipid studies results were not recorded and there is no data on what lipid or BP treatment was given. The number of subjects studied was not large. Finally, there was no funding. Nevertheless, this analysis finds that age normalized CIMT quartiles predicted total mortality.

We did not exclude those subjects with a prior CVA or TIA because most CVAs and TIAs are not related to atherosclerosis. Specifically, ∼40% of CVAs and TIAs are caused by hypertension and ∼40% are caused by atrial fibrillation while only ∼10% of CVAs and TIAs are caused by carotid atherosclerosis. This is borne out by the finding of analysis of 0.9m people that SBP and age predict CVA (10), but the atherosclerotic risk factors of total cholesterol and HDL cholesterol did not predict CVA (1).

## THE NOVEL LIFE TABLE MODEL

Cox proportional hazard models (6, 7) are excellent statistical tools for most clinical trials. If the hazards are high as with aggressive cancers or heart failure, then age is less important since the disease rather than age is driving the outcome. Also, if cohorts of similar aged subjects are studied (as in some prospective epidemiological studies), then Cox proportional hazard models can be appropriate.

In the case where starting age is more variable, where subjects enter at varying ages and the hazards from the studied disease are complicated by other age-related diseases, then age should be considered when performing statistics, especially when a trial is prolonged into ages with higher mortality (such as the 8^th^ and 9^th^ decade in Fig. 2). For these cases, this paper proposed two life table models. The Known life table model did not include those subjects lost to follow-up (similar to traditional Cox model). This model will lose statistical power given the elimination of those lost to follow-up.

The ALL life table model included all of the subjects including those lost to follow-up. Including those lost to follow-up could contaminate some statistical models because those lost to follow-up are more likely to have had an outcome that is not measured (7). Some authors (8) suggest that subjects lost to follow-up can be sometimes be included. The proposed All life table model is likely to be such a model because it includes known vital status *at each age* for each subject. In the All model, data ends in the life table for both those lost to follow-up and those alive at the end of the trial. As such, the All model is less contaminated by outcomes occurring either after the trial has ended or when a subject is lost to follow-up since there are no entries for either in the life table. If both the ALL and Known model show similar results, then there will be more confidence in the result.

Interestingly, the both the All and Known life table models both tended to find more statistical difference than the Cox model when the cohort had a wide age difference as in this study.

Importantly, the life table model reports not only significance, but a clearly understandable measure of effect size: age difference (AD) can be easily understood by those not fully trained as statisticians.

## Data Availability

All data produced in the present study are available upon reasonable request to the authors

## Acknowledgements

The author would like to thank Professor Carlos Ayers (deceased) who was instrumental in the creation of the CIMT program at the University of Virginia.

